# Differential Regulation of ERK and mTOR Signaling Pathways in Failing Human Hearts

**DOI:** 10.1101/2020.12.17.20248326

**Authors:** Rebecca Autenried, Eric T. Weatherford, Yuan Zhang, Helena C. Kenny, Renata O. Pereira, Brian T. O’Neill, Patrick Ten Eyck, Kenneth C. Bedi, Kenneth B. Margulies, E. Dale Abel

## Abstract

**Objectives:** We hypothesized that disruption of pathways downstream of insulin signaling characterize pathological ventricular remodeling and may provide insights into the pathophysiology of heart failure. To test this hypothesis, we examined components of the insulin signaling pathway in tissue explants from human hearts obtained from healthy donors and explants from heart failure patients with and without diabetes, receiving a heart transplant.

**Background:** Pathologic ventricular remodeling accompanied by hypertrophic growth is a common characteristic of heart failure including in patients with diabetes. The contribution of aberrant insulin signaling in the pathophysiology of diabetes-associated heart failure and, ventricular hypertrophy is incompletely understood.

**Methods:** Hearts of twenty non-failing donor participants and thirty-one human cardiac transplant patients were assessed for insulin signaling. Samples were sorted into four groups: non-failing non-obese (NFN), non-failing obese (NFO), failing non-diabetic (FND), and failing diabetic (FDM). Ejection fraction was assessed by echocardiography and clinically relevant systolic dysfunction was defined as left ventricular ejection fraction <50%. A clinical diabetes diagnosis was obtained from chart review. As a proxy measure of prolonged glycemia, plasma fructosamine was determined by colorimetric assay. Insulin signaling, protein phosphorylation, and total protein levels were measured by immunoblot.

**Results:** When all groups were analyzed together, hyperglycemia correlated with increased cardiac size and decreased function. Cardiac size correlated with increased levels of insulin receptor (IRb) and phosphorylated ERK but with decreased levels of phosphorylated Akt and mTOR. IRb and p-Akt correlated with fructosamine, but p-ERK and p-mTOR did not. Cardiac hypertrophy correlated with decreased GLUT1 levels, increased Hexokinase I and repression mitochondrial complexes I, III and IV in concert with activation of AMPK.

**Conclusions:** Altered insulin signaling, characterized by increased IRb content, activation of ERK but repression of Akt and mTOR signaling pathways is present in the end-stage failing human heart. Similar divergence of insulin signaling pathways have been previously described in vascular smooth muscle.

**CONDENSED ABSTRACT:** We hypothesized that disruption of pathways downstream of insulin signaling characterize pathological ventricular remodeling and may provide insights into pathophysiology. To test this hypothesis, we examined components of the insulin signaling pathway in tissue explants from human hearts obtained from healthy donors and explants from heart failure patients with and without diabetes, receiving a heart transplant. We found that altered insulin signaling, characterized by increased IRb content and activation of ERK but repression of Akt and mTOR signaling pathways is present in the end-stage failing human heart.

**HIGHLIGHTS:** In this cross-sectional analysis of end-stage failing human cardiac tissue, hyperglycemia correlated with cardiac dysfunction and increased cardiac hypertrophy.

1. While myocardial insulin resistance may exist in the PI3K-Akt-mTOR pathway in end-stage failing human hearts, ERK signaling is induced, which may contribute to cardiac hypertrophy in a manner that is independent of plasma insulin.
2. Differential activation of branches of insulin signaling in human failing hearts, supports the concept of selective insulin resistance.
3. These findings have implications for the consequences of modulating systemic insulin sensitivity in patients with heart failure.

## 1. INTRODUCTION

The risk of heart failure increases four-fold with type-I diabetes and two-fold with type II diabetes [1]. Chronic hyperglycemia and insulin resistance may adversely influence left ventricular remodeling, predisposing patients with diabetes to developing heart failure [2]. Although it is well-known that failing hearts manifest altered insulin signaling, the interactions between hyperglycemia and myocardial insulin signaling in the myocardium of adults with end-stage heart failure are incompletely understood. Furthermore, few studies have comprehensively investigated insulin signaling pathways in a cohort of human cardiac tissue explants. This study investigated changes in the insulin signaling pathway in failing hearts from diabetic and non-diabetic patients and compared these to non-failing hearts in lean and obese non-diabetic donors.

### 2. METHODS

### 2.1 Human Study Participants

Heart tissue samples were collected from twenty, non-failing donor participants, who were stratified into lean (NFL) and obese (NFO) groups based on body mass index. Thirty-one other subjects, who were receiving cardiac transplants for end-stage heart failure, were stratified into non-diabetic (FND) and diabetic (FDM) groups, based on a clinical diagnosis from patient’s chart review (Figure 1A). All study procedures were approved by the University of Pennsylvania Hospital Institutional Review Board, and prospective informed consent for research use of heart tissue was obtained from all transplant recipients and next-of-kin in the case of organ donors. Cardiac- and metabolism-related prescriptions and past medical history were obtained from chart review (Supplemental Table 1).

**Figure 1.**
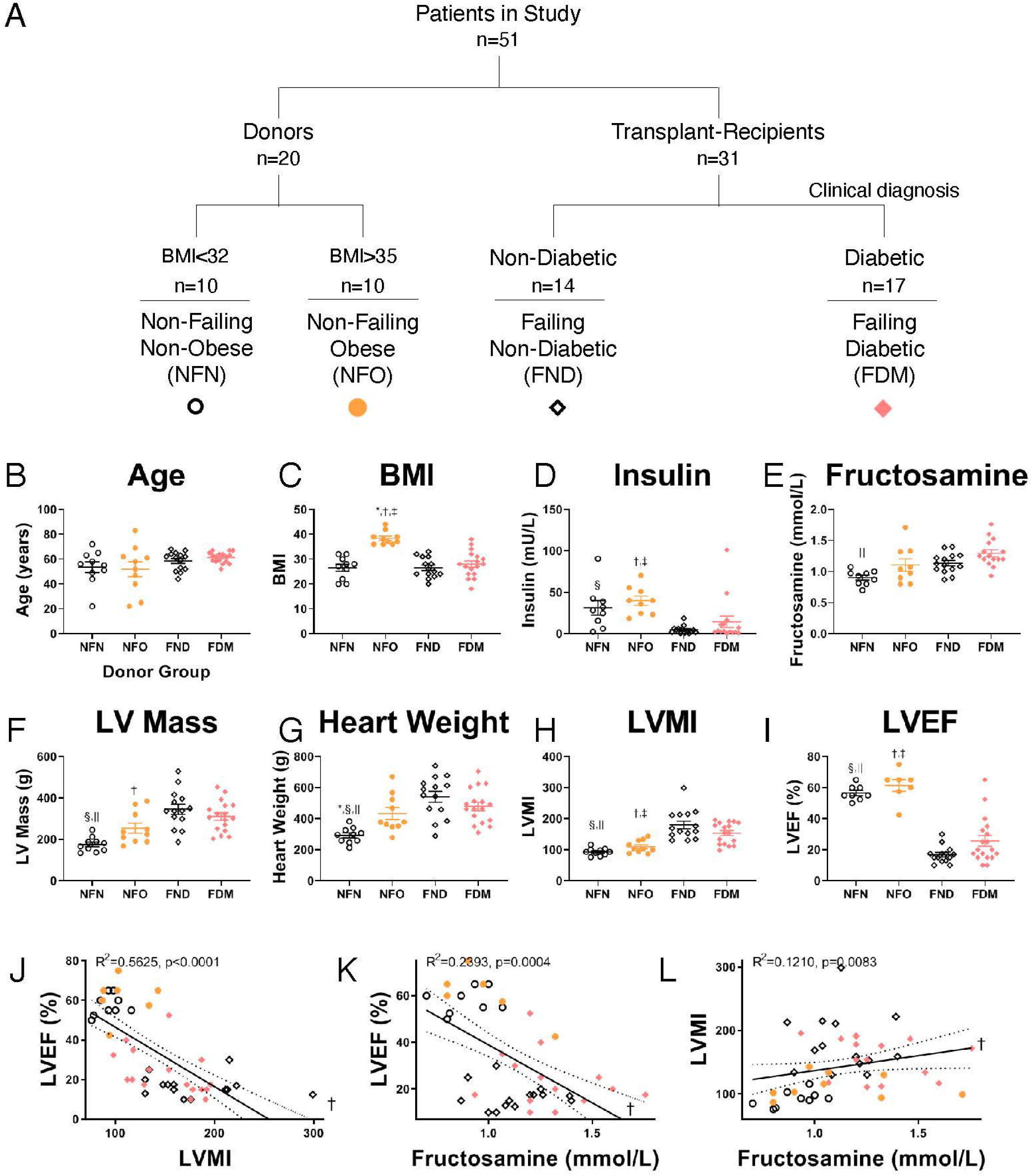
Hyperglycemia correlates with decreased cardiac function and cardiac hypertrophy. Flowchart of study participants and key to graph icons (A). Age and BMI (B-C), plasma concentrations of insulin and fructosamine (D-E) Heart weight and function (F, H, I) from echocardiography or determined at time of heart removal (G). All study values are plotted with the mean ± SEM. ANOVA p<0.05 indicated with * NFN v NFO, † NFO v FND, ‡ NFO v FDM, § NFN v FND, || NFN v FDM. Correlation graphs of ventricular mass and function in relation to fructosamine (J-L) with regressions ± 10% CI and R^2^ and p-values depicted. † p<0.05 by Spearman ranked correlation.

### 2.2 Cardiac Tissue Collection

Procurement of whole human hearts was performed under protocols and ethical regulations approved by Institutional Review Boards at the University of Pennsylvania and the Gift-of-Life Donor Program (Pennsylvania, USA). Non-failing hearts were obtained at the time of organ donation from cadaveric donors. Failing human hearts were procured at the time of orthotropic heart transplantation at the Hospital of the University of Pennsylvania following informed consent from all participants. In all cases, hearts were arrested *in situ* using ice-cold cardioplegia solution and were transported in 4°C Krebs-Henseleit Buffer on wet ice. Transmural left ventricular samples, excluding epicardial fat to the greatest extent possible, were dissected from the mid-left ventricular free wall below the papillary muscle. Left ventricular tissues were flash frozen in liquid nitrogen within 4□h of explantation, and stored at −80 °C.

### 2.3 Cardiac Function

Ejection fraction was assessed by echocardiography and clinically relevant systolic dysfunction was defined as left ventricular ejection fraction less than fifty percent. Complete echocardiographic data was not available on all of the donor patients, but some data, e.g. ejection fraction, was obtained during evaluation for cardiac transplant. Cardiac geometric parameters – including heart weight, left ventricular myocardial index (LVMI), left ventricular end diastolic diameter (LVEDD), left ventricular end systolic diameter (LVESD), and posterior wall thickness – were calculated from echocardiographic data (Figure 1F-I and Supplemental Figure 1D-L).

### 2.4 Plasma Measurements

Plasma was obtained at the time of organ transplant or tissue collection. All patients are fasted for a minimum of 12 hours with donors being fasted for a minimum of 24 hours. Glucose was measured with a colorimetric detection kit using 2µl of plasma (Invitrogen, Carlsbad, CA). Normal glucose range is 60–110 mg/dL, [3]. Insulin levels were quantified using an Insulin ELISA from 25µl of plasma (Crystal Chem, Elk Grove Village, IL). As a proxy measure of glycemic status, plasma fructosamine was determined by colorimetric assay from 50µl of plasma (Kamiya Biomedical, Tukwila, WA) [4].

### 2.5 Protein Levels and Activation

For immunoblotting, ∼10 mg of frozen tissue was homogenized in 600 µl RIPA lysis buffer using steel beads and a bead-beater (Qiagen TissueLyser II). RIPA buffer contained: 150 mM sodium chloride, 1.0% NP-40, 0.5% sodium deoxycholate, 0.1% SDS (sodium dodecyl sulfate), and 50 mM Tris, pH 8.0. Gel acrylamide percentage was optimized for each protein of interest. Tissue lysates were resolved on SDS-PAGE and transferred to nitrocellulose membranes. Primary antibodies used are summarized in Supplemental Table 2. Antibodies were tested on blots with positive control samples from cell lysates to ensure the specificity of proteins of interest. IRDye 800CW anti-Mouse (LICOR, Lincoln, NE) and Alexa fluor anti-Rabbit 680 (Invitrogen, Carlsbad, CA) were used as secondary antibodies, and fluorescence was quantified using the LICOR Odyssey imager (LICOR, Lincoln, NE). Image Studio Lite Version 5.2 software was used to quantify the densitometry of western blot bands. Because there were too many samples in the study to fit on one SDS-PAGE gel, two gels were run in parallel and treated exactly the same for every step (Supplemental Figures 2, 3, and 4). To ensure that quantifications were accurate when pooling data from both membranes, histone 3 (H3) was selected as a loading control as it did not correlate with variables examined.

### 2.6 Statistical Analysis

Data is presented as scatter plots with bars to represent the means ± the standard error of the mean. Western blot results are presented as ratios of phosphorylated to total protein levels. Total protein levels were normalized to Ponceau S staining as a measure of total protein loaded per lane. Column statistics were run for each group. The D’Agostino & Pearson and Shapiro-Wilk normality tests were used to determine whether the data was normally distributed. If so, a t-test was performed, if not, non-parametric analysis was applied using the Mann-Whitney test. A probability value of p≤0.05 was considered significantly different. One-way ANOVA was used to analyze normally distributed four-group (non-failing lean, non-failing obese, failing non-diabetic, and failing diabetic) comparisons, and the Kruskal-Wallis was used for non-normally distributed comparisons. Correlations and linear regressions were determined for the left ventricular immunoblot data versus plasma fructosamine, left ventricular ejection fraction, left ventricular myocardial index, and plasma insulin datasets following outlier removal using the ROUT method. If datasets were normally distributed, Pearson and Spearman correlations were calculated for normally and non-normally distributed measures, respectively. One-tailed p values less than 0.05 were considered statistically significant. The line of best fit was plotted as a solid line with 95% confidence intervals plotted as dotted lines. Statistical calculations were performed using Microsoft Excel and the GraphPad Prism software. We also used SAS 9.4.

To further determine if clinical variables could confound the relationships between signaling proteins and cardiac structure and function, additional statistical analyses were performed. Initial associations were made between each pair of main measures using Pearson’s unadjusted and partial correlations. Partial correlations were then calculated, adjusting for Body Mass Index (BMI), sex, HTN, pacemaker, insulin, plasma insulin, and oral hypoglycemics, individually. Plots were generated displaying the relationship between unadjusted and partial correlations for each variable pairing to visually identify which are noticeably modified by one of the adjustment variables. Points were stratified by group (FDM, FND, NFN, NFO) to further visually discern if there is a group effect as well. Correlations with a p-value < 0.05 are considered statistically significant.

## 3. RESULTS

### 3.1 General Characterization of Study Population

This study examined explanted hearts obtained from subjects with end-stage heart failure with reduced ejection fraction and compared them with un-transplanted donor hearts. The study population was further divided into four groups based on BMI or a clinical diagnosis of diabetes (Figure 1A). The four study groups were age-matched (Figure 1B), and a BMI of 32 was used to segregate non-failing groups with or without morbid obesity (Figure 1C). Additionally, the failing groups were segregated by the presence or absence of a clinical diagnosis of diabetes. For a general characterization of our study population, plasma insulin levels were highest in the Non-Failing Obese (NFO) and unexpectedly lowest in the failing groups (Figure 1D). Fructosamine reflects average blood glucose levels over a two-week period and relative to non-failing non-obese (NFN) was significantly elevated in the NFO, FND, and FDM groups (Figure 1E). Left Ventricular Mass, Heart Weight, and Left Ventricular Mass Index (LVMI) were increased in the failing as compared to the non-failing groups (Figure 1F-H). The NFO had elevated weight and body surface area (BSA, Supplemental Figure 1 A and C) but no difference in height across the study groups (Supplemental Figure 1B). Thus, although heart weight was increased in the NFO group, these differences were lost when LVMI, which accounts for body surface area, was calculated (Figure 1F and H). Ejection fractions for the FND and FDM groups are shown in Figure 1I. Prescribed medications, past medical and surgical history are summarized in Supplemental Table 1. As expected, LV Ejection Fraction (LVEF) correlated negatively with increasing LVMI in the study population (Figure 1J). LVEF correlated negatively while LVMI correlated positively with increasing plasma fructosamine (Figure 1K-L).

Percent of left ventricle divided by heart weight and heart mass index were elevated in the failing groups (Supplemental Figure 1D and E). Left ventricular end diastolic and systolic diameters were elevated in the failing groups (Supplemental Figure 1F and G). Posterior wall thickness is reduced in the FDM as compared with the FND (Supplemental Figure 1H). There were no statistically significant differences between the FND and FDM in cardiac measurements of right atrial pressure, pulmonary artery systolic, diastolic, pulmonary capillary wedge pressure, and cardiac index (Supplemental Figure 1I-M). Additionally, blood creatinine and plasma glucose and triglycerides did not show a difference between the four study groups (Supplemental Figure 1N-P).

### 3.2 ERK, Akt, and mTOR Signaling in Failing and Non-failing Human Hearts

Correlation analyses were performed on multiple signaling parameters with measures of cardiac structure, function, or glycemia (see Figure 6A for a diagram of how the insulin signaling related proteins relate to one another within the cardiomyocyte). Insulin receptor (IR) and phosphorylated AKT (pAkt) levels were not different across each of the four study groups (Figure 2A, E). However, in the entire study population, protein levels of insulin receptor in the myocardium correlated positively with LVMI (IRb, R^2^=0.1918, p=0.0015), negatively with LVEF (R^2^=0.1586, p=0.0050), and positively with fructosamine (R^2^=0.05232, p=0.0779) (Figure 2B, C, and D). Next we examined kinases implicated in the regulation of ventricular size and cardiac hypertrophy. Phosphorylation of Akt correlated negatively with LVMI (R^2^=0.0975, p=0.063), positively with LVEF (R^2^=0.1227, p=0.0106), and negatively with fructosamine (R^2^=0.2111, p=0.0010) (Figure 2F, G, and H). Interestingly, ERK phosphorylation was decreased in the NFO study group relative to the other three groups (Figure 2I) but correlated positively with LVMI (R^2^=0.1771, p=0.0020), negatively with LVEF (R^2^=0.2178, p=0.0010), and did not correlate with fructosamine (Figure 2J, K, L). mTOR phosphorylation was equivalent in the non-failing groups however mTOR phosphorylation was significantly higher in the NFN group relative to both heart failure groups (Figure 2M). Phospho-mTOR correlated negatively with LVMI (R^2^=0.0963, p=0.0169), positively with LVEF (R^2^=0.1514, p=0.0050), and did not correlate with fructosamine (Figure 2N, O, P). Additional signaling proteins were investigated. Phospho-AS160 was elevated in the NFO group as compared with the others groups (Supplemental Figure 2A) while showing no correlation with LVMI, LVEF, or fructosamine in the entire study population (Supplemental Figure 2B, C, and D). Phospho-protein phosphatase 2A (PP2A) was largely not different between the study groups (Supplemental Figure 2E) but correlated negatively with LVMI (R^2^=0.1558, p=0.0030), positively with LVEF (R^2^=0.1041, p=0.0174), and negatively with fructosamine (R^2^=0.0474, p=0.0802, Supplemental Figure 2F, G, and H). STAT3 phosphorylation was elevated in the NFO as compared with the FND (Supplemental Figure 2I) and population-wide correlated negatively with LVEF (R^2^=0.1966, p=0.0013, Supplemental Figure 2K). Phosphodiesterase (PDE)4 correlated negatively with LVEF (R^2^=0.0680, p=0.0437) and positively with fructosamine (R^2^=0.1055, p=0.0157, Supplemental Figure 2O and P).

**Figure 2.**
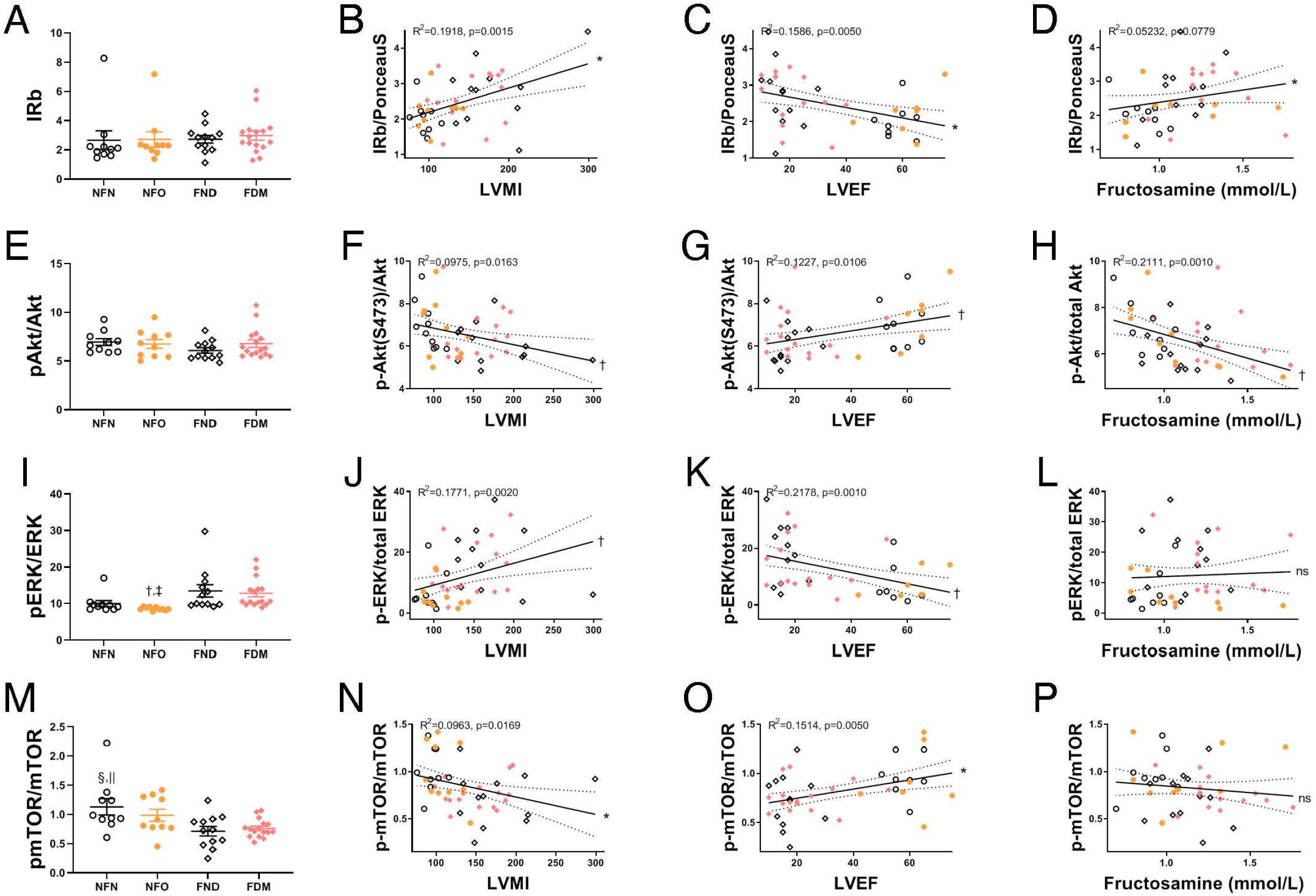
ERK, Akt and mTOR Signaling in Failing Human Hearts. Absolute protein levels in the four clinical groups (A, E, I, M); ANOVA p<0.05 indicated with, † NFO v FND, ‡ NFO v FDM, § NFN v FND, || NFN v FDM. Correlations of signaling pathways with cardiac size (LVMI, B, F, J, N), function (LVEF, C, G, K, O), and fructosamine (D,H, L, P). Graphs represent all patients in the study with regressions (± 10% CIs), correlation coefficients, and p-values as indicated. Significance indicated by * for Pearson and † for Spearman correlation coefficients.

### 3.3 Relationships Between Cardiac Size, Function, and Glycemia, with Glucose Metabolism Proteins

Given recent evidence linking altered metabolism and heart failure[5], we examined proteins linked to glucose metabolism with measures of cardiac structure, function and glycemia. In the entire study population, GLUT1 protein levels correlated negatively with LVMI (R^2^=0.1508, p=0.0032), positively with LVEF (R^2^=0.2650, p=0.0002) and negatively with fructosamine (R^2^=0.08296 p=0.0290) (Figure 3, B, C, and D). Hexokinase I (HKI) protein levels correlated positively with LVMI (R^2^=0.0966, p=0.0158), did not correlate with LVEF, and correlated positively with fructosamine (R^2^=0.08617, p=0.0266) (Figure 3F, G, and H). Hexokinase II (HKII) correlated positively with fructosamine (R^2^=0.0810, p=0.0395, Figure 3L). OGlcNac correlated negatively with LVEF (R^2^=0.0391, p=0.0991) and positively with fructosamine (R^2^=0.1222, p=0.0100, Figure 3O and P). PFKM (phosphofructokinase, muscle) correlated negatively with LVMI (R^2^=0.0708, p=0.0369, Supplemental Figure 3B). Glyceraldehyde-3-Phosphate Dehydrogenase (GAPDH) correlated negatively with fructosamine (R^2^=0.0662, p=0.0523, Supplemental Figure 3H). Pyruvate Kinase (PK)M2 correlated negatively with LVEF (R^2^=0.0588, p=0.0586) and positively with fructosamine (R^2^=0.0451, p=0.0858, Supplemental Figure 3K and L). Lactate Dehydrogenase (LDH)A correlated negatively with LVMI (R^2^=0.0737, p=0.0325) and fructosamine (R^2^=0.0420, p=0.0937, Supplemental Figure 3N and P).

**Figure 3.**
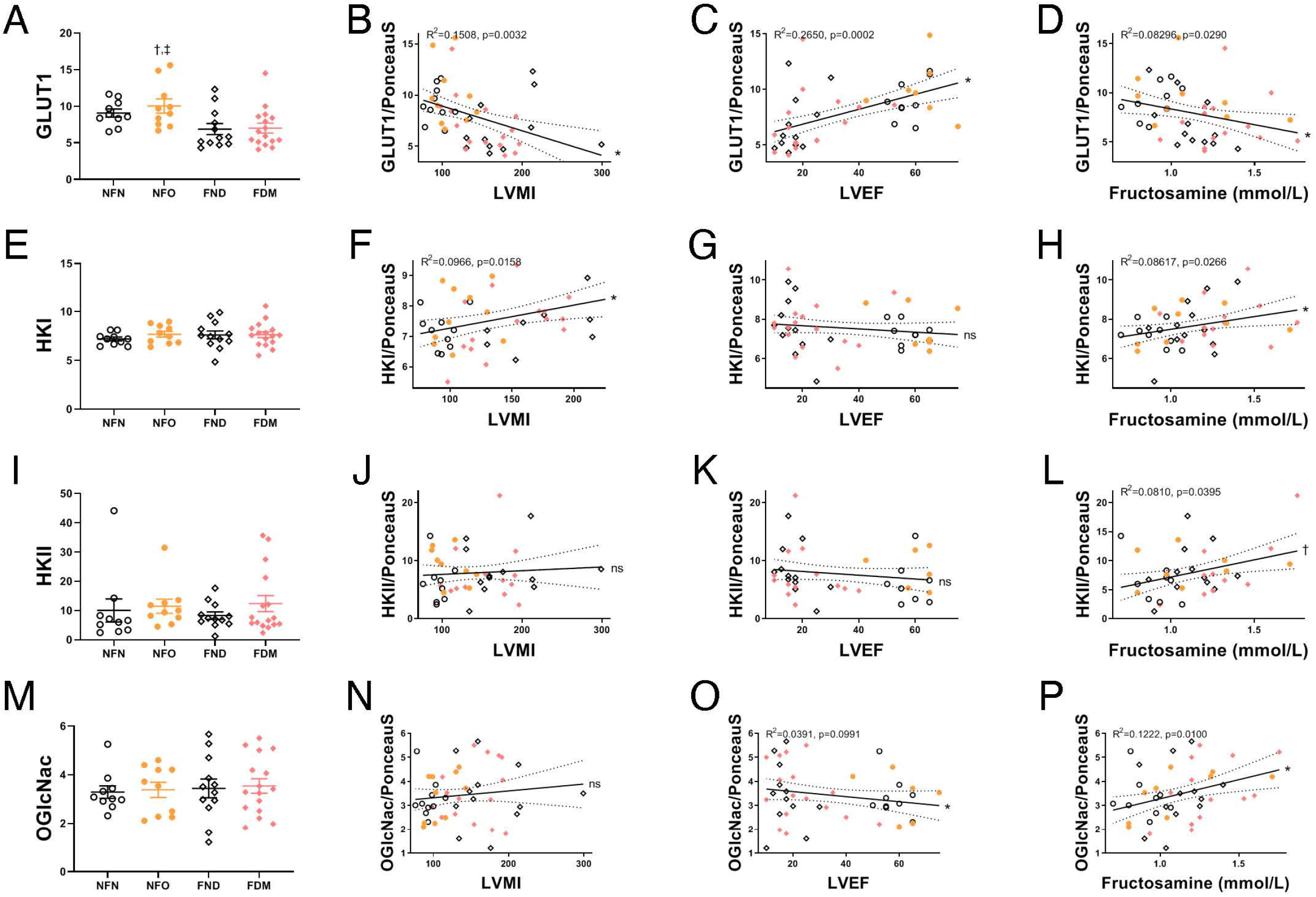
Relationships between glucose metabolism proteins, cardiac size, function, and glycemia. Absolute protein levels in the four clinical groups (A, E, I, M); ANOVA p<0.05 indicated with, † NFO v FND, ‡ NFO v FDM. Correlations of glucose metabolism related proteins with cardiac size (LVMI, B, F, J, N), function (LVEF, C, G, K, O), and fructosamine (D,H, L, P). Graphs represent all patients in the study with regressions (± 10% CIs), correlation coefficients, and p-values as indicated. Significance indicated by * for Pearson and † for Spearman correlation coefficients.

### 3.4 Relationships Between Cardiac Size, Function, and Glycemia, with AMPK and Electron Transport Chain Mitochondrial Proteins

In the entire study population, AMPK phosphorylation correlated positively with LVMI (R^2^=0.1290, p=0.0071), negatively with LVEF (R^2^=0.09400, p=0.0241) and did not correlate with fructosamine (Figure 4B, C, and D). Levels of a subunit of Complex I protein levels were significantly reduced in FDM relative to NFN (Figure 4 E) and correlated negatively with LVMI (R^2^=0.1674, p=0.0019), positively with LVEF (R^2^=0.1845 p=0.0018), and negatively with fructosamine (R^2^=0.2887, p<0.0001) (Figure 4F, G, and H). Complex II protein levels correlated positively with LVMI (R^2^=0.1249, p=0.0074) and fructosamine (R^2^=0.0647, p=0.0499) and negatively with LVEF (R^2^= 0.1300, p=0.0088, Figure 4J, L, and K). Complex III protein levels which were significantly reduced in patients with heart failure relative to the non-failing groups (Figure 4M) correlated negatively with LVMI (R^2^=0.1395, p=0.0063) and fructosamine (R^2^=0.0528, p=0.0743) and positively with LVEF (R^2^=0.3572, p<0.0001, Figure 4N, P, and O). Complex IV protein levels correlated negatively with LVMI (R^2^=0.1405, p=0.0051) and positively with LVEF (R^2^=0.1222, p=0.0116, Figure 4R and T).

**Figure 4.**
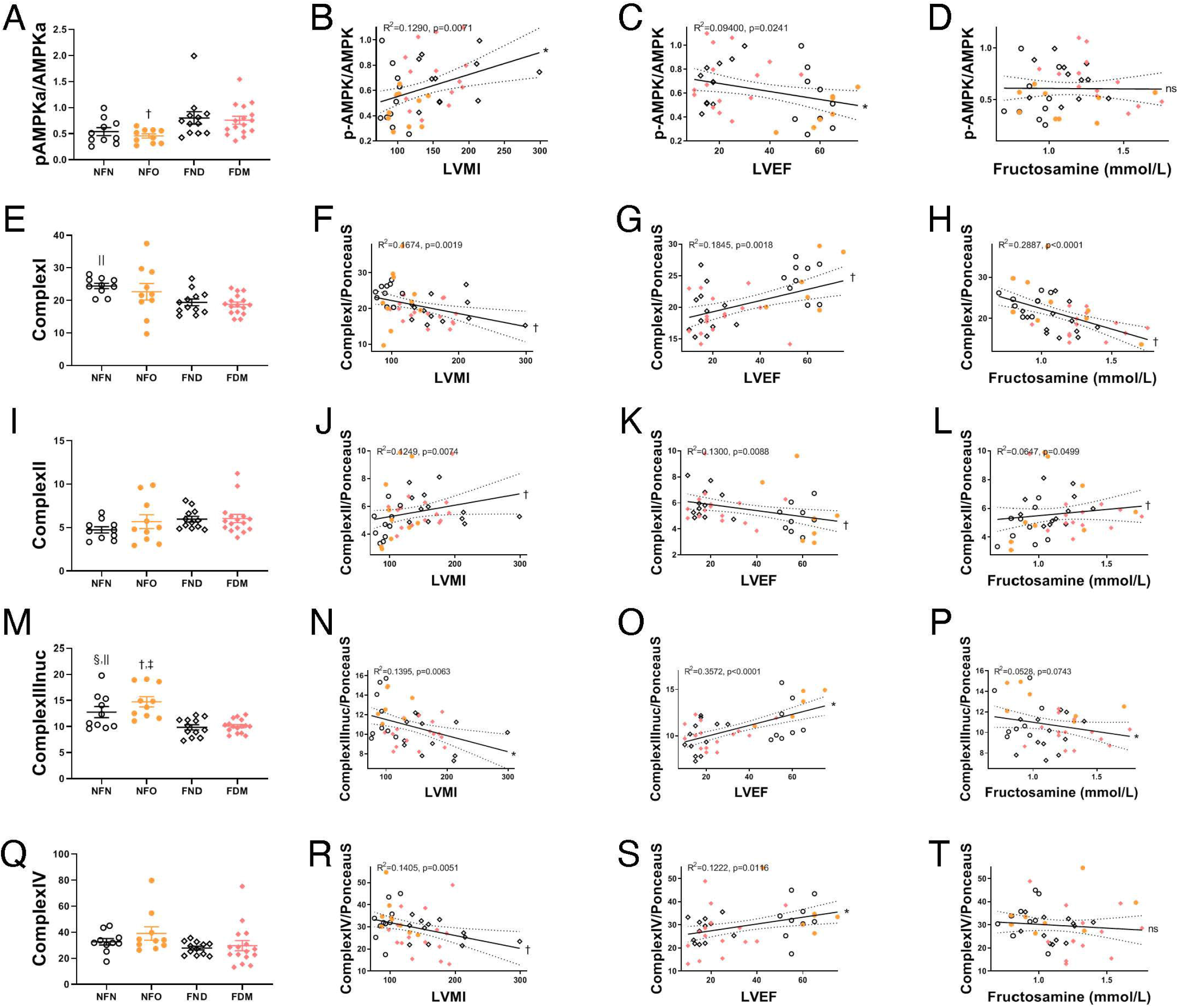
Relationships between mitochondrial proteins, cardiac size, function, and glycemia. Absolute protein levels in the four clinical groups (A, E, I, M, Q); ANOVA p<0.05 indicated with † NFO v FND, ‡ NFO v FDM, § NFN v FND, || NFN v FDM. Correlations of mitochondrial proteins with cardiac size (LVMI, B, F, J, N, R), function (LVEF, C, G, K, O, S), and fructosamine (D,H, L, P, T). Graphs represent all patients in the study with regressions (± 10% CIs), correlation coefficients, and p-values as indicated. Significance indicated by * for Pearson and † for Spearman correlation coefficients.

### 3.5 Testing for Confounding Clinical Variables of Ventricular Correlations

Using scatterplots of correlations we determined that the correlations between IRb, pAkt, pERK, pmTOR, pAMPK, GLUT1, HKI, and Complex I and Fructosamine, Heart Size, and Function were generally not confounded by the clinical variables of BMI, Sex, history of hypertension, plasma insulin, prescribed insulin, or hypoglycemics (Figure 5A-F). Outliers were noted in the NFO group for correlations adjusted to history of hypertension (Hx of Htn). These were not confounding in the molecular level analyses under consideration (Supplemental Table 3). For the FDM, FND, and NFN groups adjustment for history of hypertension did not alter the correlation of these variables (r > 0.95). In the NFO the correlations though reduced, remained statistically significant (r = 0.51, p<0.01).

**Figure 5.**
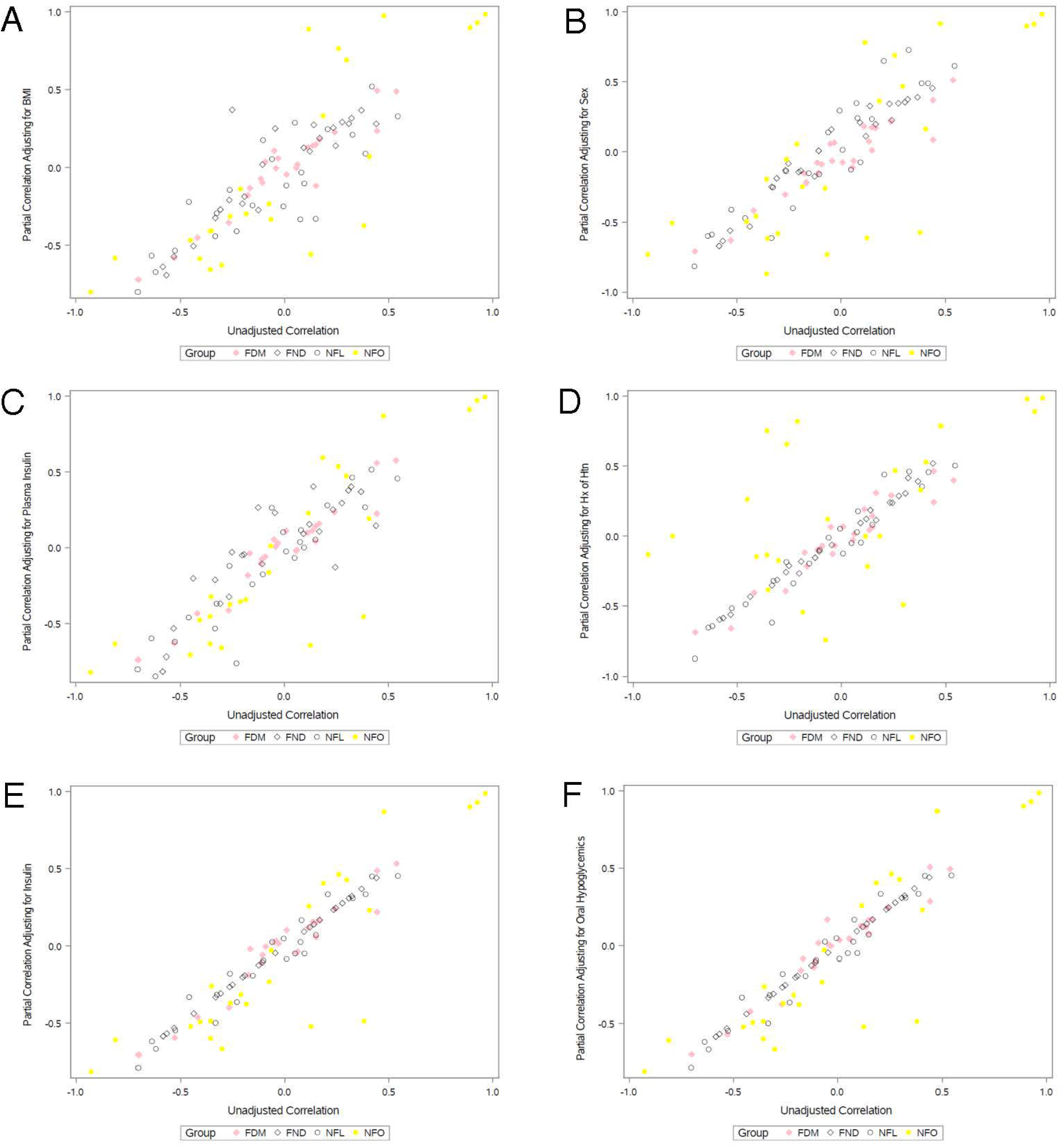
Scatterplot of Correlations. Each symbol represents the correlation between eight variables and three clinical measures leading to a total of twenty four correlates per group. The x-axes are correlations between IRb, pAkt, pERK, pmTOR, pAMPK, GLUT1, HKI, and Complex I and Fructosamine, Heart Size, and Function. The y-axes are these correlations adjusted for the axis title variable: BMI (A), Sex (B), history of hypertension (C), plasma insulin (D), prescribed insulin (E), or hypoglycemics (F). When the correlation coefficients cluster along the 45° diagonal, the relationships are not confounded by the adjusted variable.

Figure 6 summarizes the main findings of this study in schematic form.

**Figure 6.**
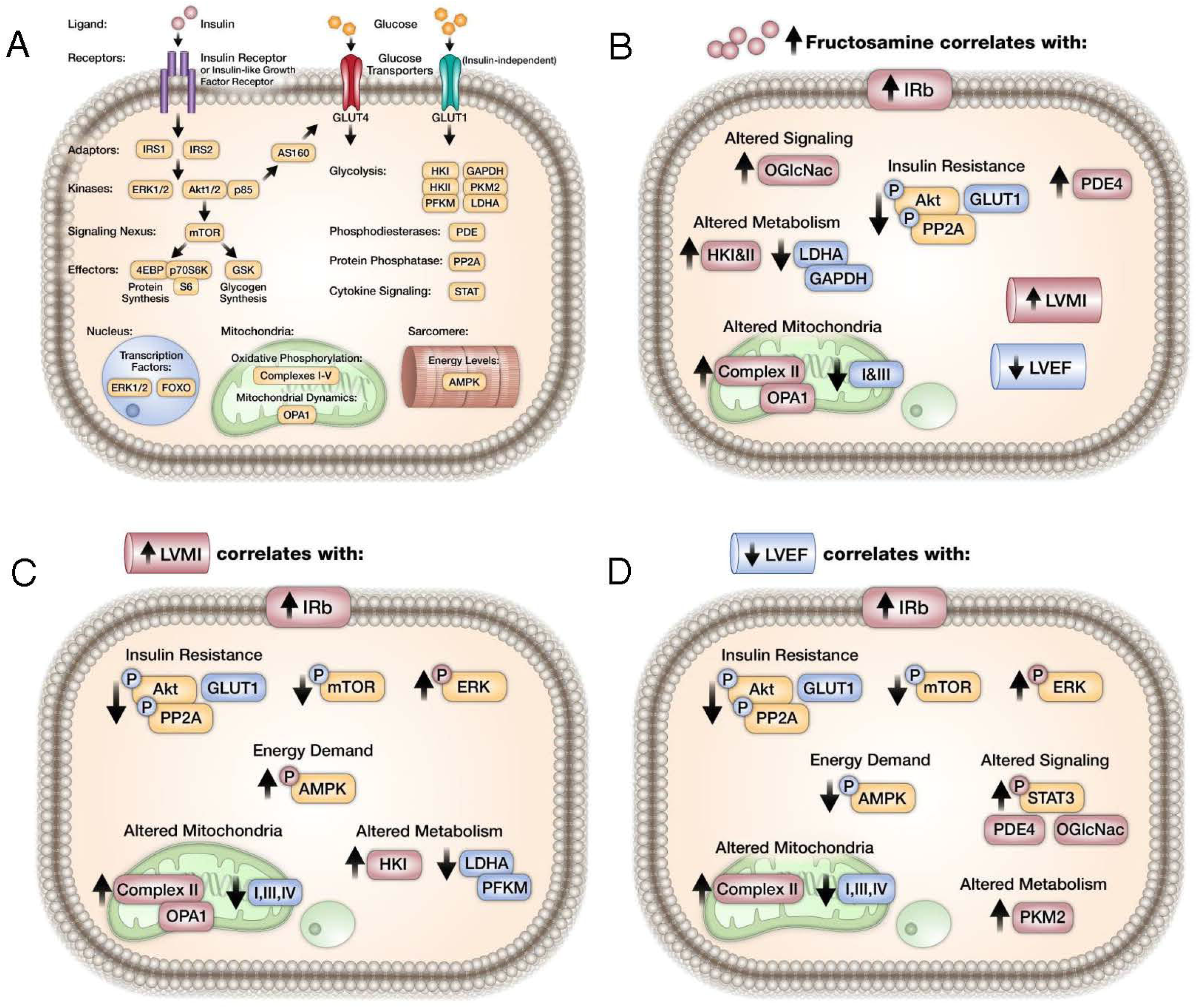
Schematic Diagram of Insulin Signaling Proteins. (A) and Summary of Study Findings (B-D). In B-D red background and blue background indicates increase or decrease respectively; P in circle indicates phosphorylated protein normalized to total protein levels.

## 4. DISCUSSION

### 4.1 Summary and significance

Systemic and myocardial insulin signaling is altered in states of cardiac dysfunction, especially in diabetes [2]. Furthermore, a significant predictor for clinical heart failure is preceding pathological hypertrophy. The aim of the present study was to investigate the relationships between cardiac structure, function, dysglycemia and insulin signaling in the left ventricle of obese human heart donors or diabetic patients with heart failure. Our results reveal that cardiac hypertrophy and dysfunction correlate with increased ERK and decreased mTOR phosphorylation in the left ventricle. The presence of increased IRb but decreased Akt activation in failing hearts, could indicate insulin resistance in these hearts. However, given that circulating insulin levels were low at the time of cardiac harvest it is likely that neither plasma insulin nor prescribed insulin significantly influenced these signaling pathways, supporting the conclusion that these signaling changes are most likely intrinsic to the heart.

### 4.2 Insulin Signaling

IR content and ERK phosphorylation correlated with indices of LV remodeling such as increased LV mass and decreased EF. In contrast, Akt and mTOR exhibited a divergent pattern. The divergent regulation of Akt and ERK has been previously described in vascular smooth muscle cells [6] and in skeletal muscle [7]. This pattern of one distal insulin signaling pathway being repressed (mTOR) and another insulin signaling pathway being activated (ERK) could indicate selective myocardial insulin resistance. In general, ERK signaling promotes cardiac hypertrophy [8] which is consistent with our findings. Furthermore, fructosamine correlates with increasing IR content and decreasing Akt phosphorylation, potentially linking dysglycemia with impaired insulin signaling to Akt in the failing myocardium. In a transgenic mouse model, knockdown of insulin receptor results in smaller hearts, which was rescued with increased expression of Akt [9]. Thus the changes in Akt activation in this study were unexpected. A negative correlation between IR and LV function supports previously described relationships between insulin receptor activation and heart failure [2] however, the pattern of correlations for Akt phosphorylation suggests insulin resistance.

Hyperphosphorylation of Akt in mouse hearts has been described following trans-aortic constriction [10]. However, Akt and mTOR in human hearts were regulated in a direction that is opposite to what could be expected if they were drivers of cardiac hypertrophy. Finally elevated PDE4 in correlation with fructosamine was negatively related to cardiac function (Supplemental Figure 2 P and O). These findings supports earlier findings by Wang, et al that revealed a mechanism linking increased signaling via insulin receptors and IRS1 with induction of PDE4 in the context of obesity related heart failure in mice and in in left atrial appendages from subjects with type 2 diabetes [11].

### 4.3 mTOR signaling

There is a large body of literature (mainly in animal models) indicating that cardiac hypertrophy correlates with mTOR activation [12]. However, it is possible that decreased Akt and mTOR activation in end-stage human heart failure could reflect de-activation of pro-survival pathways that could correlate with increased cell death. Of interest, Akt signaling inversely correlates with long term measures of glycemia. Thus, it is possible that in human hearts, Akt and mTOR activation parallels hyperglycemia in the face of systemic insulin resistance.

In mouse models, embryonic mTOR knockout revealed the necessity of mTOR for cardiomyocyte survival and proliferation [13] while inducible mTOR knockout impaired the hypertrophic response to pressure-overload leading to progressive heart failure [14]. In mouse models of pathological cardiac hypertrophy and heart failure, induced by pressure overload, mTOR inactivation was observed, whereas exercise-induced physiological hypertrophy was accompanied by activation of mTOR phosphorylation [15]. Kinase dead and constitutively active mouse models argued against a role for mTOR in cardiac size regulation, while having significant roles in signaling and cardiac function [16]. Rapamycin treatment reduced cardiac hypertrophy in response to pressure overload, but this could have been through either the mTOR or the ERK pathway as both were less activated in the treated mice [17]. Our results support the hypothesis raised by McMullen et. al in mouse models, that compensated hypertrophy might involve mTOR pathway activation and that mTOR independent pathways become activated in the transition to failure [18].

### 4.4 Glucose Metabolism

GLUT1 correlated negatively with increasing LV mass and declining cardiac function while there was no change in GLUT4. In contrast, there was a divergent pattern of GLUT1 versus HKI in relation to LV remodeling. OGlcNac correlated with fructosamine, suggesting that the declining GLUT1 did not limit glucose entry into these failing hearts. Decreased GLUT4/GLUT1 ratio was observed in cardiac tissue of humans with hypertrophy in concert with reduced glucose metabolism in heart failure [19]. GLUT4 knockdown transgenic mice exhibit increased GLUT1 expression, increased myocardial glucose utilization, and increased cardiac hypertrophy [9]. Thus the GLUT1 correlations contrast with earlier studies, mainly in preclinical models describing GLUT1 induction in pathological cardiac hypertrophy or heart failure [20]. Importantly, GLUT4 quantitatively accounts for more cardiac glucose uptake than GLUT1. The increase in HKI could be consistent with an increase in glycolytic flux. Increased OGlcNac modifications reflecting increased fructosamine generation via the hexosamine biosynthetic pathway also supports increased intracellular availability of glucose or glycolytic intermediates in failing hearts [21].

### 4.5 Mitochondria

Cardiac hypertrophy and heart failure also correlated with increased AMPK phosphorylation, which is consistent with impaired bioenergetics in the failing heart. With the exception of complex II, mitochondrial ETC complexes declined with LV remodeling. Complex I is tightly correlated with glycemic control, and is regulated in a divergent manner from complex II. Heart failure is globally associated with altered mitochondrial structure and function with extensive evidence in animal models and some evidence in human studies [22]. Our findings support these prior studies. The divergent regulation by complex II is intriguing and the mechanisms responsible for its differential regulation relative to other ETC subunits is incompletely understood. It is interesting to speculate if imbalanced stoichiometry of ETC subunits could contribute to mitochondrial ROS overproduction in the failing human heart.

### 4.6 Limitations of the Study

The main limitation of this study is the relatively small sample size. Furthermore, even though the study groups were relatively well age-matched, the circumstances at organ collection were significantly different between the non-failing accidental deaths and the explanted failing hearts of patients receiving cardiac transplants. It should also be noted that within the failing groups there were more males than females, which could potentially bias these groups towards larger heart weight. This potentially confounding factor is most likely adjusted for by the left ventricular myocardial index (LVMI) metric which normalizes heart weight to body surface area. Furthermore, the implications of differences in circulating insulin concentrations at the time of tissue harvest and the contribution of the neurohumoral state at the time of tissue harvest is not easily resolved. It is possible that ambient glucose concentrations at the time of explantation could have influenced some of the signaling changes observed in this study. However, because multiple factors could influence random glucose concentrations, we believe that fructosamine more closely reflects ambient circulating glucose concentrations over the antecedent 2-week period. Studies were performed in whole heart tissue which includes cardiomyocytes, fibroblasts, and endothelial cells. Thus, we cannot discern which cell type is predominantly driving any signaling change that is observed. Finally, while these studies are by their nature correlative they have generated hypotheses that could be tested in heart failure disease models. Moreover, the insights gained identify potential signaling pathways that could participate in heart failure disease progression and may reveal potential targets that could be therapeutically targeted.

### 4.7 Overall conclusions, possible implications, and future directions

In this cross-sectional analysis, hyperglycemia correlated with cardiac dysfunction and increased cardiac hypertrophy. Differential activation of branches of the insulin signaling pathway in human failing hearts, supports the concept of selective insulin resistance. The signaling changes studied, although part of classical insulin signaling pathways, might not necessarily indicate changes in insulin signaling per se, as other signaling pathways could influence these molecules as well. Taken together, while myocardial insulin resistance may exist in the PI3K-Akt-mTOR (4EBP, GSK3b, and AS160) pathway in end-stage failing human hearts, ERK signaling is induced, which may contribute to cardiac hypertrophy in a manner that is independent of plasma insulin. These findings have implications for the consequences of modulating systemic insulin sensitivity in patients with heart failure. For example, diabetic patients with heart failure treated with the insulin sensitizer metformin have beneficial effects on cardiac function [23] and more recently SGLT2 inhibitors have revealed dramatic effects on reducing the risk of cardiac failure in individuals with diabetes and in those with heart failure in the absence of diabetes [24, 25]. Increased knowledge of the role of insulin signaling in diabetic patients with heart failure could increase our understanding of mechanisms linking these novel therapies with the management of both conditions.

## Data Availability

Original data is available on request. These data will be available through the graduate student's dissertation deposited at the University of Iowa, Iowa City, IA 52246.

## ACKNOWLEDGMENTS

We thank Drs. Chris Adams, Isabella Grumbach, Matt Potthoff, and James Ankrum for critical review of this manuscript. We thank Rhonda Souvenir, Young-Do Koo, Marcelo Correia, Antentor J. Hinton Jr., Satya Tadinada, Quanjiang Zhang, Jennifer Streeter, Dao-Fu Dai, and Pablo Morales Campos for helpful discussions. We thank Nicholas Borcherding and Knute Carter for statistical support and Teresa Ruggle for summary illustrations. We thank Katharina Heinrich, Michael Kegel, and Kelsi Dahms for technical support and assay development. We thank the University of Iowa Medical Scientist Training Program, the Department of Molecular and Cellular Biology, and the Divisions of Endocrinology & Metabolism and Cardiology in Internal Medicine for administrative support.

