## Supplemental Figures for "Differential Regulation of ERK and mTOR Signaling Pathways in Failing Human Hearts"

### SUPPLEMENT FIGURES

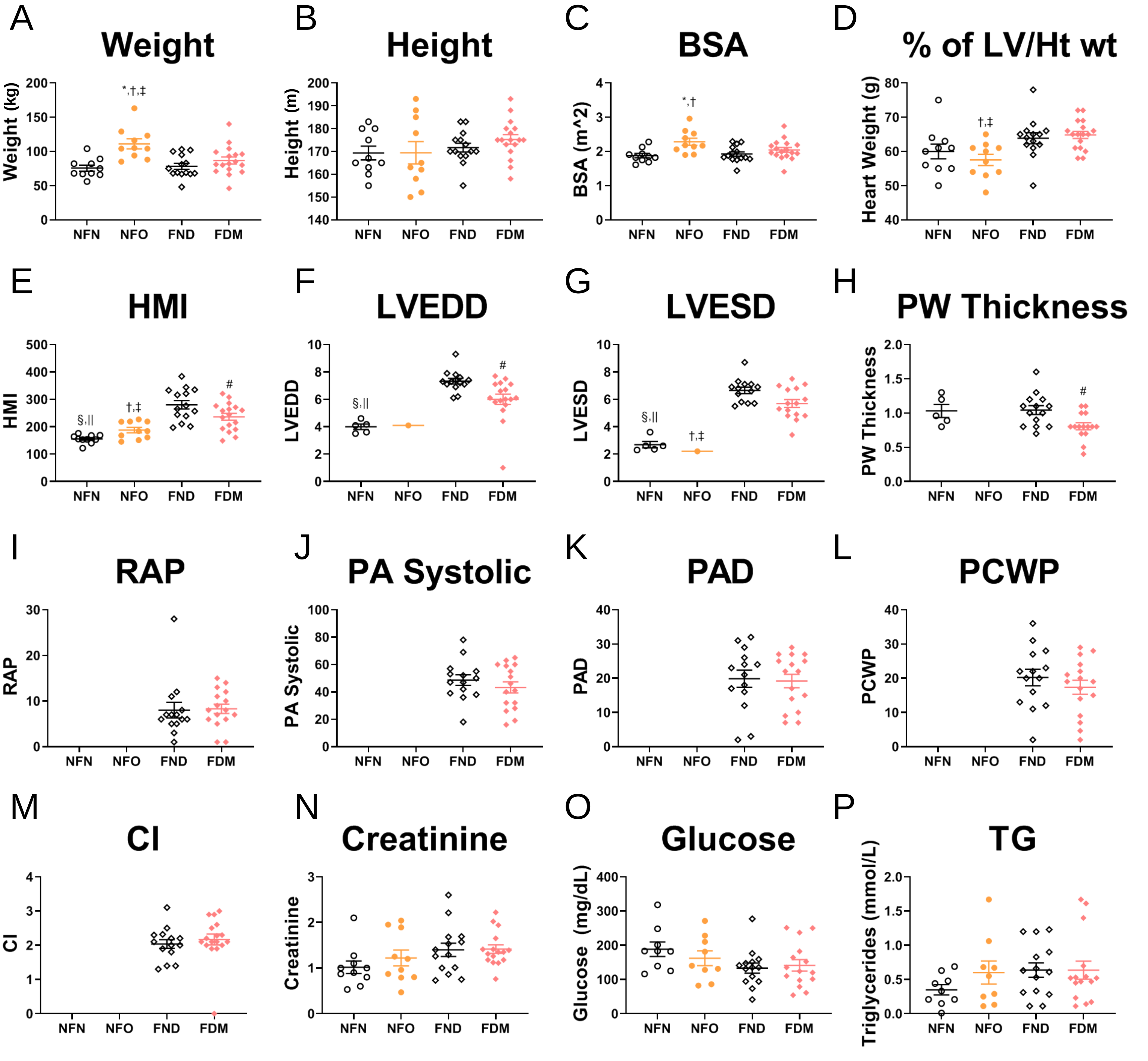


Supplemental Figure 1. **Selected clinical and laboratory data**. Scatter plots ± SEM, * NFN v NFO, † NFO v FND, ‡ NFO v FDM, § NFN v FND, || NFN v FDM, # FND v FDM. Age, weight, height, body surface are (BSA), and body mass index (BMI) are from chart review (A-E). Left ventricular (LV) mass, heart weight, % of LV/Ht wt, heart mass index (HMI, heart weight divided by body surface area), left ventricular mass index (LVMI), left ventricular end diastolic diameter (LVEDD), left ventricular end systolic diameter (LVESD), posterior wall (PW) thickness, and left ventricular ejection fraction (LVEF) are from echocardiography (F-N). Not all calculations were available for the non-failing groups (K-M). Right atrial pressure (RAP), pulmonary artery (PA) systolic, pulmonary artery diastolic (PAD), pulmonary capillary wedge pressure (PCWP), and cardiac index (CI) are calculated from wedge pressures (O-S) and are only available for the failing groups. Plasma creatinine was obtained from chart review (T). Plasma glucose, insulin, fructosamine, and triglycerides (TG) were measured using commercially available assays (U-X).

| | NFL (%) | NFO (%) | FND (%) | FDM (%) | PVAL4 | PVAL2 | PVAL3 |
|---|---|---|---|---|---|---|---|
| Insulin Rx | 0 | 0 | 0 | 29 | 0.0156 | 0.1429 | 0.1904 |
| Hypoglycemics Rx | 0 | 0 | 0 | 41 | 0.0005 | 0.0338 | 0.1123 |
| Hx of DM | 0 | 0 | 0 | 100 | <.0001 | <.0001 | 0.0001 |
| Hx of HF | 0 | 0 | 100 | 100 | <.0001 | <.0001 | <.0001 |
| Hx of Htn | 40 | 60 | 86 | 76 | 0.0994 | 0.0314 | 0.0563 |
| Hx of ChrAfib | 0 | 20 | 86 | 41 | <.0001 | 0.0004 | 0.0003 |
| Hx of VT/VF | 0 | 0 | 71 | 47 | <.0001 | <.0001 | <.0001 |
| Hx of MI | 0 | 0 | 43 | 47 | 0.0029 | 0.0003 | 0.0011 |
| ASA Rx | 20 | 30 | 50 | 82 | 0.0053 | 0.0042 | 0.0107 |
| Lipid Lowering Rx | 30 | 10 | 64 | 65 | 0.0141 | 0.0036 | 0.0043 |
| Thyroid med Rx | 10 | 20 | 0 | 29 | 0.1304 | 1 | 1 |
| Beta Blocker Rx | 0 | 30 | 100 | 94 | <.0001 | <.0001 | <.0001 |
| ACE I Rx | 0 | 20 | 64 | 65 | 0.0008 | 0.0001 | 0.0002 |
| Angio II Antag Rx | 0 | 0 | 29 | 29 | 0.0698 | 0.008 | 0.0331 |
| Hydralazine Rx | 0 | 0 | 0 | 41 | 0.0005 | 0.0338 | 0.1123 |
| Ca Chan Blocker Rx | 0 | 0 | 14 | 0 | 0.142 | 0.5137 | 1 |
| Nitrate Rx | 0 | 0 | 36 | 29 | 0.0321 | 0.0039 | 0.0178 |
| Dig Rx | 0 | 0 | 71 | 41 | <.0001 | <.0001 | 0.0001 |
| Diuretic Rx | 0 | 10 | 93 | 94 | <.0001 | <.0001 | <.0001 |
| Amiodarone Rx | 0 | 0 | 50 | 35 | 0.0038 | 0.0006 | 0.0038 |
| Other Antiarrhythmic Rx | 0 | 0 | 36 | 0 | 0.0011 | 0.1429 | 0.1904 |
| Anti Platelet Rx | 0 | 0 | 29 | 18 | 0.1143 | 0.0338 | 0.1123 |
| Anticoagulant Rx | 0 | 10 | 64 | 35 | 0.0021 | 0.0015 | 0.0033 |
| Milirone Rx | 0 | 0 | 92 | 63 | <.0001 | <.0001 | <.0001 |
| Dobutimine Rx | 0 | 0 | 27 | 0 | 0.0266 | 0.2683 | 0.7467 |
| Prior AICD | 0 | 0 | 79 | 71 | <.0001 | <.0001 | <.0001 |
| Prior Pacemaker | 0 | 10 | 43 | 6 | 0.017 | 0.1273 | 0.3215 |
| Prior Bi V Pacer | 0 | 0 | 79 | 65 | <.0001 | <.0001 | <.0001 |
| Prior MV Repair | 0 | 0 | 0 | 12 | 0.3271 | 0.5137 | 1 |
| Prior CABG | 0 | 0 | 14 | 24 | 0.2105 | 0.0697 | 0.2024 |
| Prior Angioplasty | 0 | 0 | 14 | 12 | 0.5741 | 0.1453 | 0.4542 |
| Prior Stents | 0 | 0 | 36 | 35 | 0.0221 | 0.0035 | 0.01 |
| Prior LVAD | 0 | 0 | 0 | 0 | n/a | n/a | n/a |

Supplemental Table 1. **Medications and Medical and Surgical History**. Prior prescription medications (Rx) and medical (Hx) or surgical history (Prior) were obtained from chart review. Hypertension (htn), chronic atrial fibrillation (ChrAfib), ventricular tachycardia/ventricular fibrillation (VT/VF), myocardial infarction (MI), acetylsalicylic acid (aspirin, ASA), angiotensin converting enzyme inhibitor (ACE I), angiotensin II antagonist (Angio II Antag), Digitonin (Dig), Automatic Implantable Cardioverter Defibrillator (AICD), bi-ventricular (Bi-V) pacer, mitral valve (MV) repair, coronary artery bypass grafting (CABG), left ventricular assist device (LVAD). ANOVA PVAL4 is between all four groups, PVAL2 is between failing and non-failing, and PVAL3 is between NFL and the three groups with cardiac hypertrophy (NFO, FND, FDM).

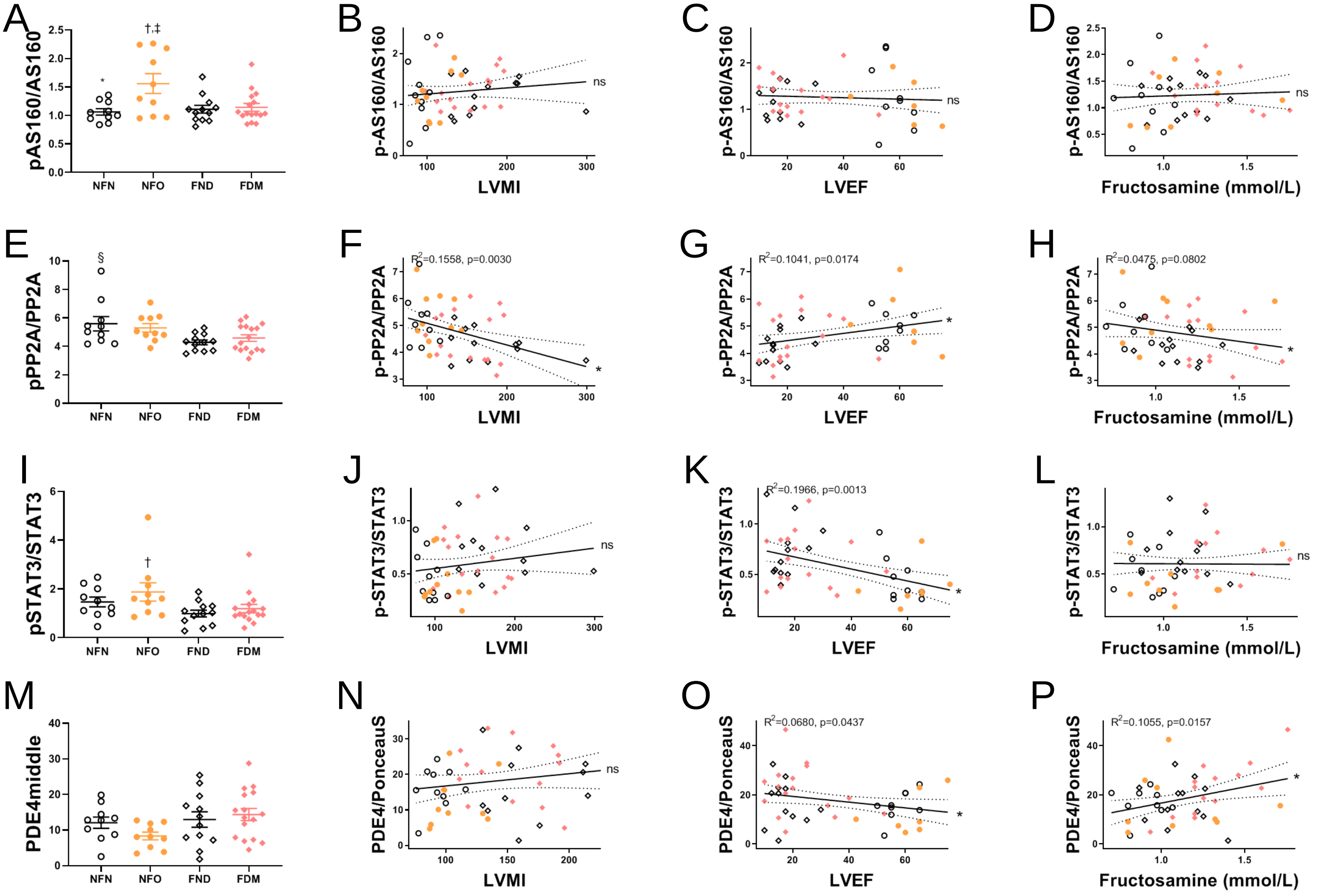


Supplemental Figure 2. **Additional correlations between signaling proteins, cardiac size, function, and glycemia**. Absolute protein levels in the four clinical groups (A, E, I, M); ANOVA p<0.05 indicated with * NFN v NFO, † NFO v FND, ‡ NFO v FDM, § NFN v FND, || NFN v FDM. Correlations of signaling pathways with cardiac size (LVMI, B, F, J, N), function (LVEF, C, G, K, O), and fructosamine (D,H, L, P). Graphs represent all patients in the study with regressions (± 10% CIs), correlation coefficients, and p-values as indicated. Significance indicated by * for Pearson and † for Spearman.

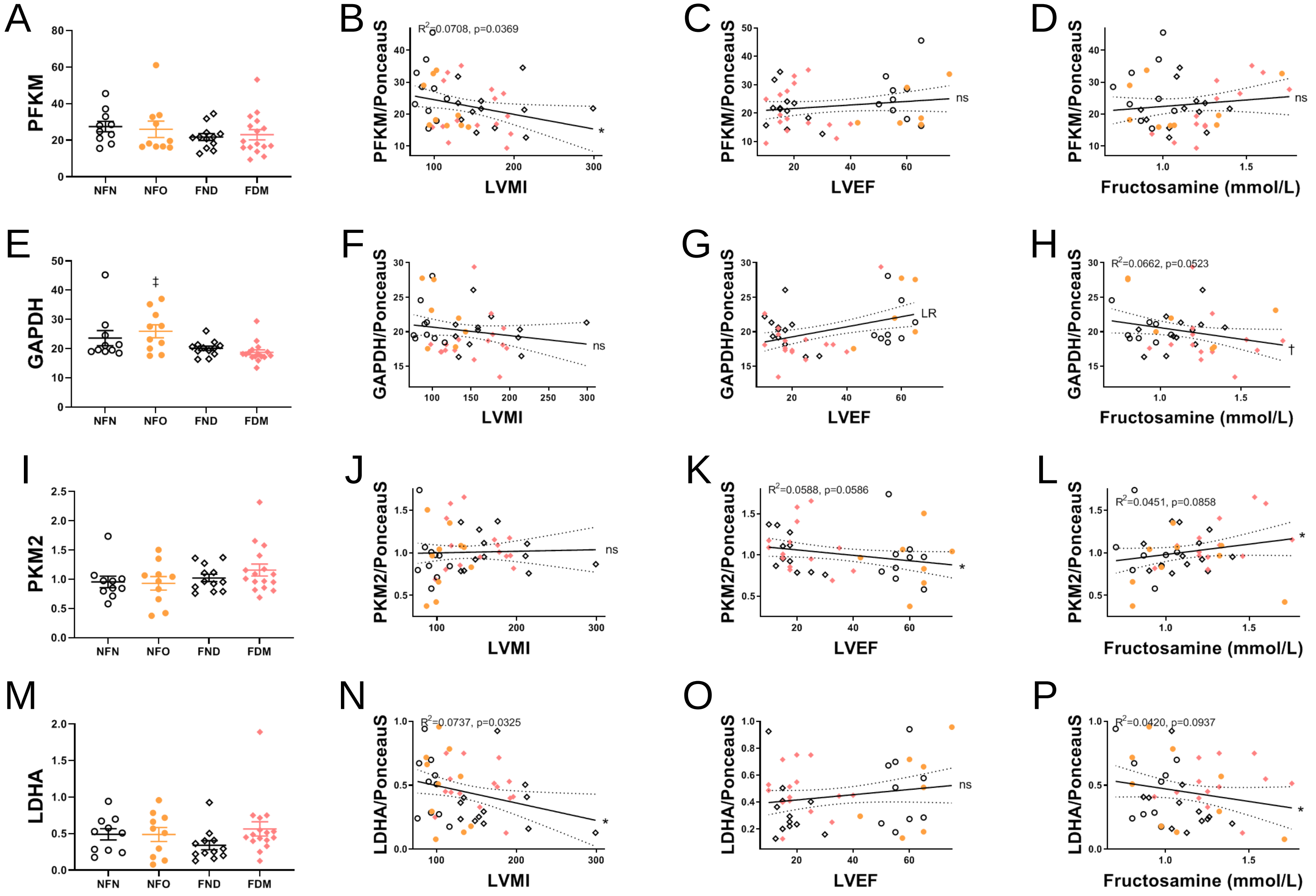


Supplemental Figure 3. **Additional correlations between metabolic proteins, cardiac size, function, and glycemia**. Absolute protein levels in the four clinical groups (A, E, I, M); ANOVA p<0.05 indicated with * NFN v NFO, † NFO v FND, ‡ NFO v FDM, § NFN v FND, || NFN v FDM. Correlations of signaling pathways with cardiac size (LVMI, B, F, J, N), function (LVEF, C, G, K, O), and fructosamine (D,H, L, P). Graphs represent all patients in the study with regressions (± 10% CIs), correlation coefficients, and p-values as indicated. Significance indicated by * for Pearson and † for Spearman.

| Protein | Company | Catalog No. |
|---|---|---|
| IRb | Cell Signaling (CS) | 3025S |
| IGFR | | 3027 |
| PI3K | | |
| p85 | | 4292 |
| p-Akt | | 9018 |
| Akt | | 2920 |
| p-Akt1 | | 9018 |
| Akt1 | | 2967 |
| p-Akt2 | | 8599 |
| Akt2 | | 5239 |
| p-ERK1/2 | | 9101 |
| ERK1/2 | | 9107 |
| p-mTOR | | 2971 |
| mTOR | | 4517 |
| p-S6 | | 2211 |
| S6 | | 2317 |
| 4EBP | | 9644 |
| p-GSK3b | | 5558 |
| GSK3b | | 9832 |
| p-GSK3a/b | | 9331 |
| p-FOXO3a | | 9465 |
| p-AS160 | | 8881 |
| AS160 | | 2670 |
| PP2A | | 2038 |
| p-AMPKa | | 2531 |
| AMPKa | | 2793 |
| p-STAT3 | | 9145 |
| STAT3 | | 9139 |
| HK1 | | 2024 |
| HK2 | | 2867 |
| PFKM | | 8164 |
| GAPDH | | 2118 |
| PKM2 | | 4053 |
| LDHA | | 3582 |
| Cytochrome c | | 4272 |
| GLUT1 | Millipore (MP) | 07-1401 |
| GLUT4 | | 07-1404 |
| PDE9 | | ABN32 |
| p-PP2A | Santa Cruz (SC) | 271903 |
| OGlcNac | Abcam (ab) | 2739 |
| panPDE4 | | 14628 |
| OPA1 | | 42364 |
| NDUFA9 | | 14713 |
| SDHA | | 14715 |
| UQCRC1 | | 110252 |
| MT-CO1 | | 14705 |
| MT-CO3 | | 110259 |
| ATP5A | | 14748 |
| H3 | | 1791 |

Supplemental Table 2. **Antibodies utilized for the left ventricular immunoblot studies**.

| 8 Variables: | None | BMI | Sex | Hx_of_Htn | Pacemaker | Insulin | PLASMA_INSULIN | Oral_Hypoglycemics |
|---|---|---|---|---|---|---|---|---|

| Simple Statistics | | | | | | | |
|---|---|---|---|---|---|---|---|
| Variable | N | Mean | Std Dev | Sum | Minimum | Maximum | Label |
| None | 96 | -0.03386 | 0.37080 | -3.25090 | -0.92888 | 0.96362 | Unadjusted Correlation |
| BMI | 96 | -0.05218 | 0.41967 | -5.00935 | -0.80099 | 0.98379 | Partial Correlation Adjusting for BMI |
| Sex | 96 | -0.02561 | 0.43716 | -2.45830 | -0.86640 | 0.98261 | Partial Correlation Adjusting for Sex |
| Hx_of_Htn | 96 | 0.0006703 | 0.40481 | 0.06435 | -0.87327 | 0.98693 | Partial Correlation Adjusting for Hx of Htn |
| Pacemaker | 96 | -0.05755 | 0.40459 | -5.52477 | -0.81168 | 0.98561 | Partial Correlation Adjusting for Pacemaker |
| Insulin | 96 | -0.05671 | 0.39655 | -5.44384 | -0.81168 | 0.98561 | Partial Correlation Adjusting for Insulin |
| PLASMA_INSULIN | 96 | -0.05297 | 0.42834 | -5.08497 | -0.84350 | 0.99254 | Partial Correlation Adjusting for Plasma Insulin |
| Oral_Hypoglycemics | 96 | -0.05385 | 0.39634 | -5.16972 | -0.81168 | 0.98561 | Partial Correlation Adjusting for Oral Hypoglycemics |

| Pearson Correlation Coefficients, N = 96<br>Prob > \|r\| under H0: Rho=0 | | | | | | | | |
|---|---|---|---|---|---|---|---|---|
| | None | BMI | Sex | Hx_of_Htn | Pacemaker | Insulin | PLASMA_INSULIN | Oral_Hypoglycemics |
| None<br>Unadjusted Correlation | 1.00000 | 0.85262<br><.0001 | 0.85587<br><.0001 | 0.75833<br><.0001 | 0.90890<br><.0001 | 0.92578<br><.0001 | 0.88714<br><.0001 | 0.92659<br><.0001 |
| BMI<br>Partial Correlation Adjusting for BMI | 0.85262<br><.0001 | 1.00000 | 0.90945<br><.0001 | 0.67892<br><.0001 | 0.91446<br><.0001 | 0.93006<br><.0001 | 0.91293<br><.0001 | 0.93071<br><.0001 |
| Sex<br>Partial Correlation Adjusting for Sex | 0.85587<br><.0001 | 0.90945<br><.0001 | 1.00000 | 0.74700<br><.0001 | 0.91148<br><.0001 | 0.92951<br><.0001 | 0.90675<br><.0001 | 0.92846<br><.0001 |
| Hx_of_Htn<br>Partial Correlation Adjusting for Hx of Htn | 0.75833<br><.0001 | 0.67892<br><.0001 | 0.74700<br><.0001 | 1.00000 | 0.72035<br><.0001 | 0.72798<br><.0001 | 0.68784<br><.0001 | 0.72792<br><.0001 |
| Pacemaker<br>Partial Correlation Adjusting for Pacemaker | 0.90890<br><.0001 | 0.91446<br><.0001 | 0.91148<br><.0001 | 0.72035<br><.0001 | 1.00000 | 0.98155<br><.0001 | 0.94475<br><.0001 | 0.98282<br><.0001 |
| Insulin<br>Partial Correlation Adjusting for Insulin | 0.92578<br><.0001 | 0.93006<br><.0001 | 0.92951<br><.0001 | 0.72798<br><.0001 | 0.98155<br><.0001 | 1.00000 | 0.96437<br><.0001 | 0.99804<br><.0001 |
| PLASMA_INSULIN<br>Partial Correlation Adjusting for Plasma Insulin | 0.88714<br><.0001 | 0.91293<br><.0001 | 0.90675<br><.0001 | 0.68784<br><.0001 | 0.94475<br><.0001 | 0.96437<br><.0001 | 1.00000 | 0.96311<br><.0001 |
| Oral_Hypoglycemics<br>Partial Correlation Adjusting for Oral Hypoglycemics | 0.92659<br><.0001 | 0.93071<br><.0001 | 0.92846<br><.0001 | 0.72792<br><.0001 | 0.98282<br><.0001 | 0.99804<br><.0001 | 0.96311<br><.0001 | 1.00000 |

Group=NFL

| Pearson Correlation Coefficients, N = 24<br>Prob > \|r\| under H0: Rho=0 | | |
|---|---|---|
| | None | Hx_of_Htn |
| None<br>Unadjusted Correlation | 1.00000 | 0.96488<br><.0001 |
| Hx_of_Htn<br>Partial Correlation Adjusting for Hx of Htn | 0.96488<br><.0001 | 1.00000 |

Group=NFO

| Pearson Correlation Coefficients, N = 24<br>Prob > \|r\| under H0: Rho=0 | | |
|---|---|---|
| | None | Hx_of_Htn |
| None<br>Unadjusted Correlation | 1.00000 | 0.51477<br>0.0101 |
| Hx_of_Htn<br>Partial Correlation Adjusting for Hx of Htn | 0.51477<br>0.0101 | 1.00000 |

Group=FND

| Pearson Correlation Coefficients, N = 24<br>Prob > \|r\| under H0: Rho=0 | | |
|---|---|---|
| | None | Hx_of_Htn |
| None<br>Unadjusted Correlation | 1.00000 | 0.99548<br><.0001 |
| Hx_of_Htn<br>Partial Correlation Adjusting for Hx of Htn | 0.99548<br><.0001 | 1.00000 |

Group=FDM

| Pearson Correlation Coefficients, N = 24<br>Prob > \|r\| under H0: Rho=0 | | |
|---|---|---|
| | None | Hx_of_Htn |
| None<br>Unadjusted Correlation | 1.00000 | 0.95711<br><.0001 |
| Hx_of_Htn<br>Partial Correlation Adjusting for Hx of Htn | 0.95711<br><.0001 | 1.00000 |

Supplemental Table 3. **Table of Pearson Correlation Coefficients**.

### SUPPLEMENT FIGURE LEGENDS

Supplemental Figure 1. Selected clinical and laboratory data. Scatter plots ± SEM, * NFN v NFO, † NFO v FND, ‡ NFO v FDM, § NFN v FND, || NFN v FDM, # FND v FDM. Age, weight, height, body surface are (BSA), and body mass index (BMI) are from chart review (A-E). Left ventricular (LV) mass, heart weight, % of LV/Ht wt, heart mass index (HMI), left ventricular mass index (LVMI), left ventricular end diastolic diameter (LVEDD), left ventricular end systolic diameter (LVESD), posterior wall (PW) thickness, and left ventricular ejection fraction (LVEF) are from echocardiography (F-N). Not all calculations were available for the non-failing groups (K-M). Right atrial pressure (RAP), pulmonary artery (PA) systolic, pulmonary artery diastolic (PAD), pulmonary capillary wedge pressure (PCWP), and cardiac index (CI) are calculated from wedge pressures (O-S) and are only available for the failing groups. Plasma creatinine was obtained from chart review (T). Plasma glucose, insulin, fructosamine, and triglycerides (TG) were measured using commercially available assays (U-X).

Supplemental Table 1. Medications and Medical and Surgical History. Prior prescription medications (Rx) and medical (Hx) or surgical history (Prior) were obtained from chart review. Hypertension (htn), chronic atrial fibrillation (ChrAfib), ventricular tachycardia/ventricular fibrillation (VT/VF), myocardial infarction (MI), acetylsalicylic acid (aspirin, ASA), angiotensin converting enzyme inhibitor (ACE I), angiotensin II antagonist (Angio II Antag), Digitonin (Dig), Automatic Implantable Cardioverter Defibrillator (AICD), bi-ventricular (Bi-V) pacer, mitral valve (MV) repair, coronary artery bypass grafting (CABG), left ventricular assist device (LVAD). ANOVA PVAL4 is between all four groups, PVAL2 is between failing and non-failing, and PVAL3 is between NFL and the three groups with cardiac hypertrophy (NFO, FND, FDM).

Supplemental Figure 2. Additional correlations between signaling proteins, cardiac size, function, and glycemia. Absolute protein levels in the four clinical groups (A, E, I, M); ANOVA $p<0.05$ indicated with * NFN v NFO, † NFO v FND, ‡ NFO v FDM, § NFN v FND, || NFN v FDM. Correlations of signaling pathways with cardiac size (LVMI, B, F, J, N), function (LVEF, C, G, K, O), and fructosamine (D,H, L, P). Graphs represent all patients in the study with regressions (± 10% CIs), correlation coefficients, and p-values as indicated. Significance indicated by * for Pearson and † for Spearman.

Supplemental Figure 3. Additional correlations between metabolic proteins, cardiac size, function, and glycemia. Absolute protein levels in the four clinical groups (A, E, I, M); ANOVA $p<0.05$ indicated with * NFN v NFO, † NFO v FND, ‡ NFO v FDM, § NFN v FND, || NFN v FDM. Correlations of signaling pathways with cardiac size (LVMI, B, F, J, N), function (LVEF, C, G, K, O), and fructosamine (D,H, L, P). Graphs represent all patients in the study with regressions (± 10% CIs), correlation coefficients, and p-values as indicated. Significance indicated by * for Pearson and † for Spearman.

Supplemental Table 2. Antibodies utilized for the left ventricular immunoblot studies.

Supplemental Table 3. Table of Pearson Correlation Coefficients
